# Changes in Resting-State Functional Network Specialization in Progressive Supranuclear Palsy

**DOI:** 10.64898/2026.11.14.26367857

**Authors:** Juan P. Vazquez, Anja Soldan, Hanzhang Lu, Marilyn Albert, Alexander Pantelyat

**Affiliations:** Department of Neurology, Johns Hopkins University School of Medicine, Baltimore, Maryland, USA

**Keywords:** PSP, rs-FMRI, neuroimaging, graph theory, biomarkers, functional connectivity

## Abstract

Progressive supranuclear palsy (PSP) is a neurodegenerative tauopathy characterized by heterogeneous clinical presentations and disruption of distributed neural networks. Resting-state functional MRI (rs-fMRI) provides a systems-level approach to examine intrinsic brain connectivity and may offer insights into disease-related network dysfunction. In this study, we compared rs-fMRI connectivity measures between individuals with PSP and healthy controls and explored differences across PSP clinical variants. 23 participants with probable or possible PSP and 23 age- and sex-matched healthy controls underwent structural and functional MRI. Graph theory metrics and functional connectivity measures across seven canonical resting-state networks were analyzed. Clinical severity was assessed using the PSP Rating Scale (PSPRS) and cognitive measures including the Montreal Cognitive Assessment (MoCA) and frontal assessment battery (FAB). Compared with controls, participants with PSP demonstrated significantly reduced global network modularity and segregation, indicating decreased functional network specialization. Functional connectivity was significantly reduced in multiple large-scale networks, including frontoparietal control, default mode, dorsal attention, somatomotor, and visual networks. No significant differences in graph theory metrics or network connectivity were observed between PSP clinical variants. These findings suggest that PSP is associated with widespread disruption of intrinsic functional networks, reflected by reduced modular organization and network segregation across several canonical resting-state systems while preserving local nodal clustering and global efficiency. Network-level alterations appear broadly similar across clinical variants, supporting the concept that PSP represents a distributed network disorder.

## Introduction

Progressive supranuclear palsy (PSP) is a neurodegenerative disease characterized by the pathologic accumulation of 4-repeat tau protein^1^. Classic PSP or Richardson syndrome (RS) is characterized by falls, vertical gaze limitations, and postural instability^2^. Clinical diagnosis of PSP has improved over the last several decades; however early and reliable detection remains challenging given considerable overlap in presentation with other neurodegenerative diseases^3,4^. Further, there is a relative lack of sensitivity in the current clinical and supporting radiologic diagnostic criteria, which may lead to an underestimation of PSP prevalence estimates (1-18 cases per 100,000^5^). There exists, in addition, substantial heterogeneity in presentations of PSP reflected by its defined clinical variants. The 2017 International Parkinson and Movement Disorder Society (MDS) diagnostic criteria define 8 subtypes of PSP^3^. Here, we focus on 3 of these subtypes: PSP-RS, PSP with speech/language disorders (PSP-SL), and PSP with corticobasal syndrome (PSP-CBS). PSP-RS is characterized by prominent frontal-executive dysfunction^6^, PSP-SL by nonfluent/agrammatic primary progressive aphasia and/or speech apraxia^7^, and PSP-CBS by notable limb praxis deficits, which are typically asymmetric^8^. This suggests that clinical impairments in these PSP subtypes may result from deterioration in the structure and function of disparate cortical-subcortical networks: a frontostriatal network in PSP-RS, a dominant inferior frontal-subcortical speech network in PSP-SL, and an asymmetric parietal-dorsolateral striatum network in PSP-CBS. There is, however, significant clinical overlap between PSP subtypes, indicating shared abnormalities in network organization.

These PSP syndromes currently lack effective treatments, significantly limit quality of life, and shorten lifespan. While there is a great interest in therapeutic development for these disorders, clinical trials thus far have been unsuccessful, hampered by a lack of a reliable diagnostic or prognostic *in vivo* biomarker^9^. Here, we aimed to compare resting-state functional MRI measures of connectivity between the aforementioned 3 PSP subtypes and healthy controls.

Resting-state functional MRI (rs-fMRI) measures low-frequency, spontaneous BOLD signal fluctuations that reveal large-scale intrinsic functional connectivity networks ie. spatially distributed sets of regions that show synchronous activity in the absence of tasks and recapitulate many task-evoked systems^10^. This systems-level view is well-suited for neurodegenerative phenotyping where pathology can affect distributed circuits^11,12^. In PSP, early pathology targets subcortical and brainstem structures, including the midbrain and cerebellar outflow pathways, with downstream cortical dysfunction often reflecting disconnection rather than primary cortical disease^13^. In this study, we focused on evaluating cortical network connectivity as a complementary approach to capture downstream effects of subcortical pathology, which may in turn reveal patterns of network reorganization that relate to clinical heterogeneity.

In order to characterize both global network topology and regional connectivity patterns, we employed graph theory metrics alongside canonical resting-state network analyses. Graph theory provides a mathematical framework for quantifying network organization. Graph theory measures of interest include modularity, reflecting the degree to which a network can be divided into subsystems (or modules); clustering coefficient, a measure of local connectivity between neighboring nodes; and global efficiency, which captures information integration across the network.

Prior seed-based and region-of-interest (ROI) analyses in PSP have demonstrated reduced connectivity within frontal, striatal, and cerebellar circuits^14^, while studies examining canonical network function have reported inconsistent results, with some showing reduced connectivity^13^ and others increased connectivity or relative preservation within and between cortical, subcortical, and brainstem regions^15^. Fewer studies have employed graph theory approaches to characterize whole-brain network topology in PSP^16^. Furthermore, direct comparisons of network connectivity across PSP variants are sparse, and the relationship between network-level disruption and clinical measures such as disease severity and cognitive function remains incompletely characterized.

The overarching goals of the study are to 1) compare functional connectivity and graph theory measures between PSP and healthy controls 2) examine differences in these measures across PSP subtypes and 3) explore associations between these measures and clinical severity, including motor symptoms and cognitive function. We hypothesized that participants with PSP would demonstrate global network dysfunction compared to healthy controls. We further hypothesized that network measures would be associated with clinical function in a domain-specific manner; that is, that connectivity within networks supporting cognitive function (e.g. frontoparietal control, default mode) would correlate with performance on cognitive measures, while connectivity within motor-related networks (e.g. somatomotor, dorsal attention) would correlate with motor severity as measured by the PSP rating scale (PSPRS).

## Methods

### Study Participants

A total of 23 participants with probable PSP (RS n=12, SL n=7, CBS n=5, Table 1) and imaging data were included in our analyses and assessed by author AP. 23 age and sex-matched healthy controls from the BIOCARD study were also included^17^. Inclusion criteria for PSP participants included consensus diagnosis of either probable PSP-RS, possible PSP-SL, or possible PSP-CBS, age 40-89 years, right-handedness, and use of English as the primary language. Exclusion criteria included Fazekas small vessel ischemic disease scores >1 (based on MRI T2-FLAIR sequences), inability to provide informed consent at enrollment, and presence of another clinically significant neurologic disorder. Inclusion criteria for the control participants were age 40-89 years, right handedness, use of English as the primary language, and unimpaired cognition, as per clinical consensus.

**Table 1.** Baseline demographic and clinical variables

|  | Controls (n=23) | PSP-RS (n=11) | PSP-SL (n=7) | PSP-CBS (n=5) | p-value (Controls vs. all PSP) | p-value (PSP variants) |
|---|---|---|---|---|---|---|
| Age at baseline visit, M±SD | 70.56±7.45 | 69±8.1 | 73.29±8.2 | 71±6.6 | 0.94 | 0.54 |
| Female sex, N (%) | 10 (0.38) | 3 (27%) | 4 (0.57) | 3 (0.6) | 1 | 0.34 |
| Years of Education, M±SD | 17.35±2.52 | 15.57±2.46 | 16.83 | 13.8 | **0.02** | 0.23 |
| Disease duration | - | 4.95±2.23 | 4.96±1.53 | 5±1.47 | - | 0.97 |
| PSPRS |  |  |  |  |  |  |
| Total score | - | 42.73±7.86 | 42.14±20.54 | 50±10.15 | - | 0.55 |
| History | - | 8±3.13 | 8.42±4.86 | 9.6±2.88 | - | 0.39 |
| Mentation | - | 5.72±3.41 | 5±5.1 | 5.8±0.84 | - | 0.91 |
| Bulbar | - | 2.91±1.76 | 2.57±1.99 | 3.8±1.64 | - | 0.51 |
| Ocular Motor | - | 8.36±3.07 | 8.29±3.81 | 7.6±3.13 | - | 0.91 |
| Limb Motor | - | 7.82±1.53 | 8.43±2.10 | 10.2±3.19 | - | 0.14 |
| Gait | - | 9.91±4.35 | 9.43±6.48 | 13±4.3 | - | 0.45 |
| PSP Staging System | - | 3.33±0.98 | 2.57±1.51 | 3.40±1.15 | - | 0.38 |
| FAB |  | 12.22±3.87 | 13.2±4.44 | 14.33±1.5 | - | 0.69 |
| TOLA | - | 197.8±11.97 | 172±38.41 | 185±11.6 | - | 0.11 |
| MOCA | - | 21.55±5.7 | 18.86±3.12 | 20.2±3.11 | - | 0.49 |

### Clinical Evaluation and Consensus Diagnosis

Participants were diagnosed with PSP using the 2017 MDS Diagnostic criteria applied by the study PI (AP)^3^.

PSP participants underwent further evaluation of disease severity using the PSP Rating Scale (PSPRS). PSPRS is a validated 100-point assessment that has been used as a primary outcome measure in multiple clinical trials of PSP. It has a History section obtained from the patient and care partner, a brief Mentation assessment targeting frontal-executive function, and Motor assessments evaluating bulbar, ocular, limb and gait function^18^. To assess global cognitive function, the Montreal Cognitive Assessment (MoCA) was used. The MoCA has been validated for use in middle-aged and older adults across the cognitive spectrum (i.e., unimpaired, mild cognitive impairment, and dementia)^19^.

The Frontal Assessment Battery (FAB) further evaluated frontal executive function^20,21^. The FAB was included to support clinical phenotyping and to enable analyses examining associations between executive function and network connectivity measures. Several studies have evaluated FAB in PSP, and one^22^ found significant correlations between regional MRI connectivity measures and FAB performance.

The Test of Oral and Limb Apraxia (TOLA) was designed to efficiently assess praxis (to verbal command, mimic, and pictured everyday objects) in a range of conditions, including PSP and Corticobasal syndrome^23^. It is a clinical assessment of oral and limb praxis that evaluates gestures (range of item scores 0-3, with 3 indicating intact gesture performance and 0 indicating total loss of ability to perform).

The BIOCARD control participants underwent annual diagnostic consensus procedures, performed by the JHU BIOCARD Clinical Core, to evaluate whether they are cognitively unimpaired, have mild cognitive impairment (MCI), or dementia, using a standardized protocol consistent with the National Institute on Aging (NIA) Alzheimer’s Disease Centers’ diagnostic guidelines. For additional details regarding the BIOCARD cognitive and clinical evaluation, see^24^. The syndromic diagnoses of MCI or dementia are based on the NIA/Alzheimer’s Association (AA) for MCI^25^ and dementia^26^. All BIOCARD participants included as controls in this study were cognitively unimpaired.

### MRI acquisition

All participants underwent MRI scanning on a 3T Philips Achieva scanner (Eindhoven, The Netherlands). Structural T1-weighted images were acquired using a magnetization-prepared rapid gradient echo (MPRAGE) sequence with the following parameters: TR = 6.7 ms, TE = 3.1 ms, shot interval 3000 ms, flip angle = 8°, FOV = 240x256 mm^2^, 170 slices with 1x1x1.2 mm^3^ voxels, and scan duration = 5 min 59 s.°. Resting-state BOLD data were acquired using an echo-planar imaging (EPI) sequence with the following parameters: number of slices = 48; field of view (FOV) = 212 x 212 mm^2^; voxel size = 3.3 x 3.3 x 3.3 mm^3^; TR = 3000 ms, TE = 30 ms; flip angle = 75° and scan duration = 420 s. Fluid-attenuated inversion recovery (FLAIR) images were acquired using a multi-slice fast spin-echo sequence (TR/TE/TI=11000/100/2800 ms with 1 c 1 x 2 mm^3^; number of slices=69, SENSE acceleration factor = 2.0; scan time = 3 min 18 sec).

### Resting-State fMRI Processing

The BOLD data underwent standard preprocessing steps using [SPM/FSL and in-house MATLAB scripts]. Preprocessing included slice timing correction, realignment, normalization to Montreal Neurological Institute (MNI) 152 volumetric space using the MPRAGE image, and spatial smoothing via a Gaussian filter with a full-width half-maximum of [4/6] mm. The BOLD image series then underwent detrending and bandpass filtering, retaining components within the 0.01–0.1 Hz frequency range to isolate low-frequency fluctuations. Frame-wise displacement (FD), calculated as the sum of absolute differentials of the six rigid-body head motion parameters, was used to identify and scrub volumes with FD ≥ 0.5 mm^27^. Following motion scrubbing, participants with fewer than [70/80] frames of usable data were excluded from analysis. Nuisance signals, including global, white matter, and cerebrospinal fluid signals, along with six rigid-body head motion parameters, were regressed out.

### Functional Network Generation

Data were parcellated into 114 regions of interest (ROIs) estimated in MNI 152 volumetric space, based on the parcellation developed by Yeo and colleagues^28^. Cross-correlation coefficients were calculated between each pair of ROIs and converted to z-scores using a Fisher z-transformation, yielding a 114×114 matrix of z-transformed values. To quantify network-level functional connectivity, the matrix was reduced to 7×7 by averaging the z-transformed values belonging to the same network, resulting in seven canonical networks: visual, somatomotor, dorsal attention, ventral attention (salience), limbic, frontoparietal control, and default mode. Within-network connectivity was calculated as the mean z-transformed correlation among all region pairs within each network

### Graph Theory and Network Connectivity Measures

Graph theory measures were obtained using the Brain Connectivity Toolbox^29^ and in-house MATLAB scripts. Modularity was computed for each subject based on the predefined subnetworks from the Yeo parcellation using an algorithm previously described^30^. Modularity quantifies the degree to which a network can be decomposed into mutually separate subnetworks, or modules, that are internally integrated, yet segregated from others and is computed based on positively and negatively weighted connections of the unthresholded correlation matrix^31,32^. Higher modularity values indicate greater separation between networks (and greater connectivity within networks). Clustering coefficient represents the connectedness of neighboring nodes. Here, local clustering coefficient was calculated as the proportion of edges between nodes divided by the number of possible edges between them with higher values (between 0 and 1) indicating greater neighboring node connectedness^33^. This number was then averaged across all nodes to an overall measure of local node connectedness. Lastly, global network efficiency was calculated as the average inverse shortest path length throughout all nodes in the network. Higher global efficiency reflects greater functional integration across long-range connections^34^. Separate analyses assessing the related measure of network segregation are not presented because modularity and segregation are highly correlated in this sample (r=0.97, p<0.001) and results for segregation were the same as for modularity (data not shown)

## Statistical Analyses

All statistical analyses were performed using R version 4.3.2. Between-group differences in demographic, clinical, and neuroimaging measures were assessed using independent sample t-tests for continuous variables and chi-square tests for categorical variables. To account for multiple comparisons, we applied the Benjamini-Hochberg False Discovery Rate (FDR) correction^35^ for all independent analyses. Corrected p-values <0.05 were considered statistically significant. Associations between rsFMRI measures (three graph theory metrics and functional connectivity within the 7 large-scale networks) and clinical scores (PSPRS, MoCA, TOLA, FAB total scores) were assessed using Pearson correlation coefficients as well as linear regression models adjusted for age, sex, and education in the PSP group. Statistical significance was determined after FDR correction. Specifically, we corrected for 7 tests in the analysis assessing associations of connectivity in the 7 large-scale networks with the four clinical scores and for 4 tests in the analysis of the 4 graph metrics in relation to the 4 clinical scores. We did not additionally correct for the 4 clinical measures, as they capture conceptually distinct functions, and because these analyses were meant to generate hypotheses that can be further evaluated in future studies.

## Results

In total, our study included 23 participants with probable or possible PSP (RS=11, SL=7, CBS=5) and 23 cognitively unimpaired age-and sex-matched controls. Control participants had higher years of education (17.4 years) compared to the PSP group (15.6 years), p=0.01. Among PSP subtypes, there were no significant differences in age, sex, years of education, or clinical scores (see Table 1 for participant characteristics).

## Functional Connectivity Measures

Within-network connectivity measures were significantly lower in PSP participants compared to controls, including the frontoparietal (0.083 vs. 0.135, p=0.001), default mode (0.119 vs. 0.164, p=0.006), dorsal attention (0.117 vs. 0.191, p=0.004), ventral attention (0.139 vs 0.184, p=0.02) somatomotor (0.225 vs. 0.387, p=0.001), limbic and visual networks (0.405 vs. 0.607, p=0.0009) (Figure 1). The pattern of results was the same when total cortical volume was additionally covaried in the analyses to account for global atrophy (data not shown). When comparing PSP subtypes, there was a significant difference in the ventral attention network, with PSP-RS having lower connectivity than PSP-CBS (PSP-RS=0.114, PSP-SL=0.132, PSP-CBS=0.204, F=4.4, p=0.03) though this did not survive FDR correction (p=0.18) (Supplementary table 1).

**Figure 1.**
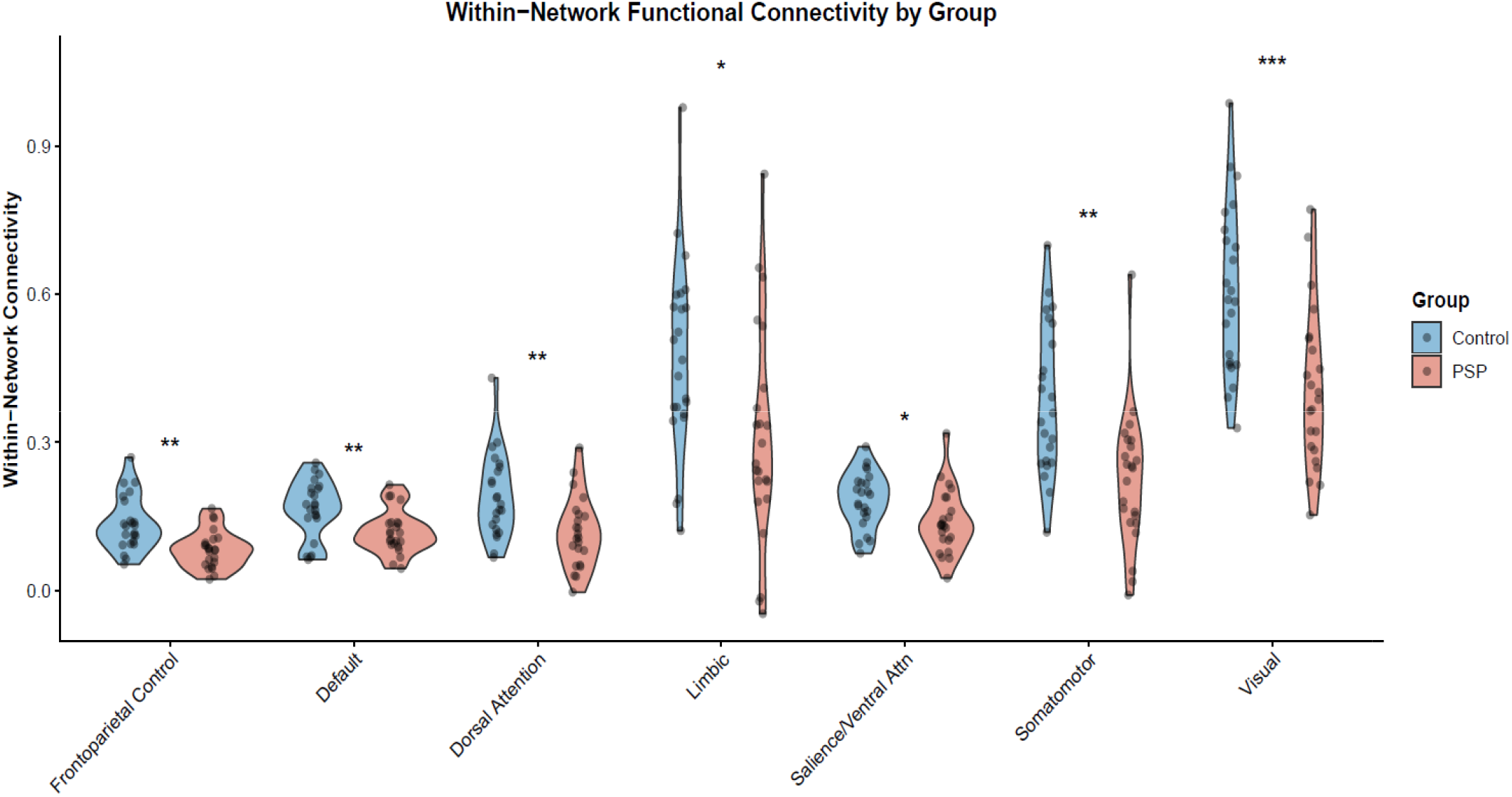
Within-network functional connectivity in PSP vs. healthy controls Within-network functional connectivity in PSP participants vs healthy controls (PSP n=23, controls n=23). Violin plots show distribution of within-network functional connectivity for seven resting-state networks where each point represents an individual participant. Asterisk indicate significant group differences. *p<0.05, **p<0.01, and ***p<0.001 FDR corrected.

We then correlated functional connectivity measures with the clinical scores of the PSP patients. Unadjusted analysis found significant positive relationships between somatomotor connectivity and PSP-RS total scores (r=0.61, p=0.002), and additionally, between dorsal attention connectivity and FAB total scores (r=0.53, p=0.028). These associations were no longer significant after FDR correction (Figure 3). When analyses were adjusted for age, sex, and years of education, a significant association was found between higher TOLA scores and somatic motor network connectivity (p=0.01); however, this did not survive FDR correction. Non-significant trends for associations, however, were seen between PSP-RS and somatic motor connectivity, FAB and dorsal attention connectivity, and TOLA and ventral attention networks (Table 2).

**Figure 2.**
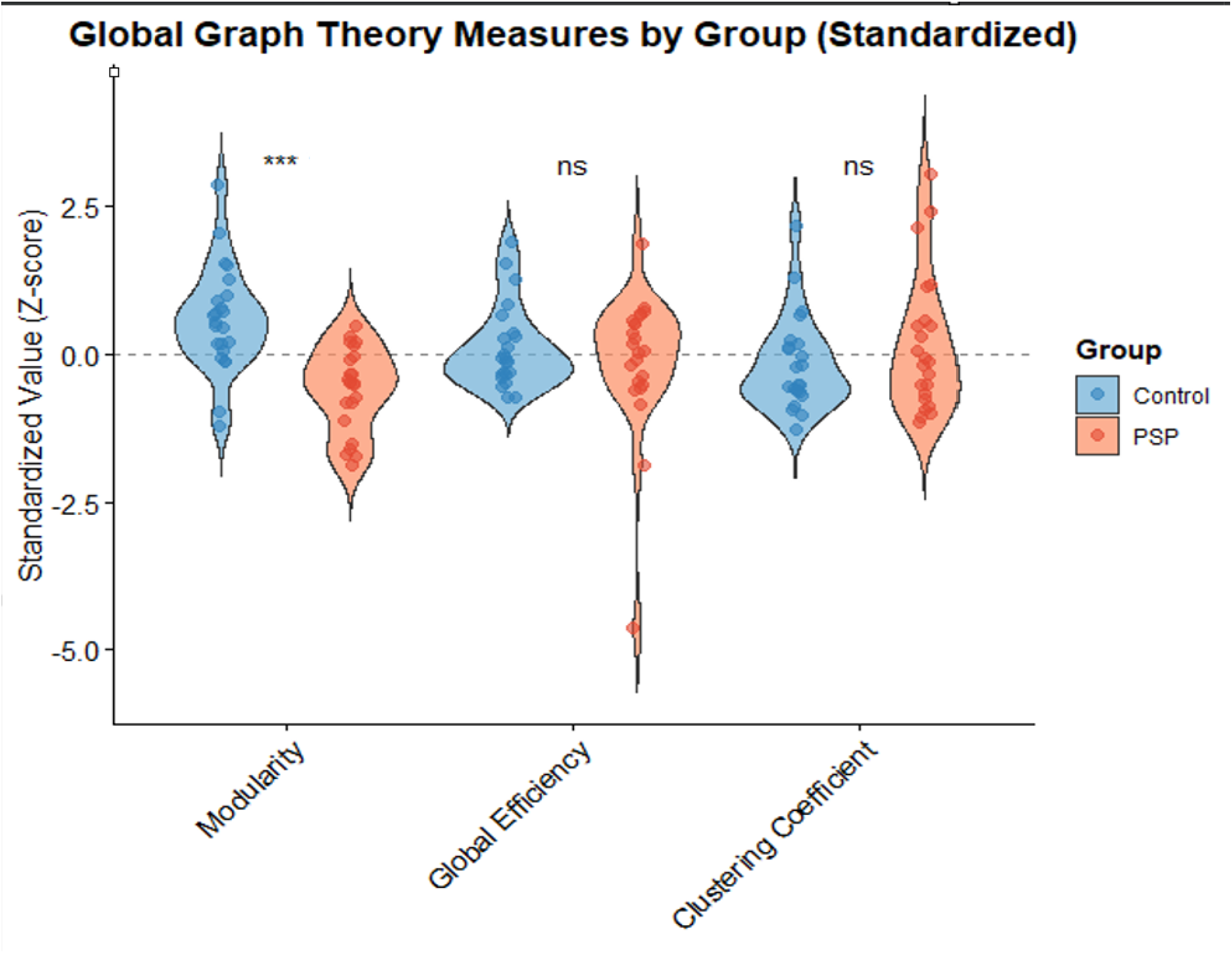
Global graph theory measures in PSP vs. healthy controls Global graph theory metrics in PSP participants vs healthy controls (PSP n=23, Controls n=23). Violin plots show distribution of standardized graph theory metrics. Values were standardized to Z-scores (mean=0, SD=1) across all participants. ***=p<0.001, FDR corrected ns=no significance

**Figure 3.**
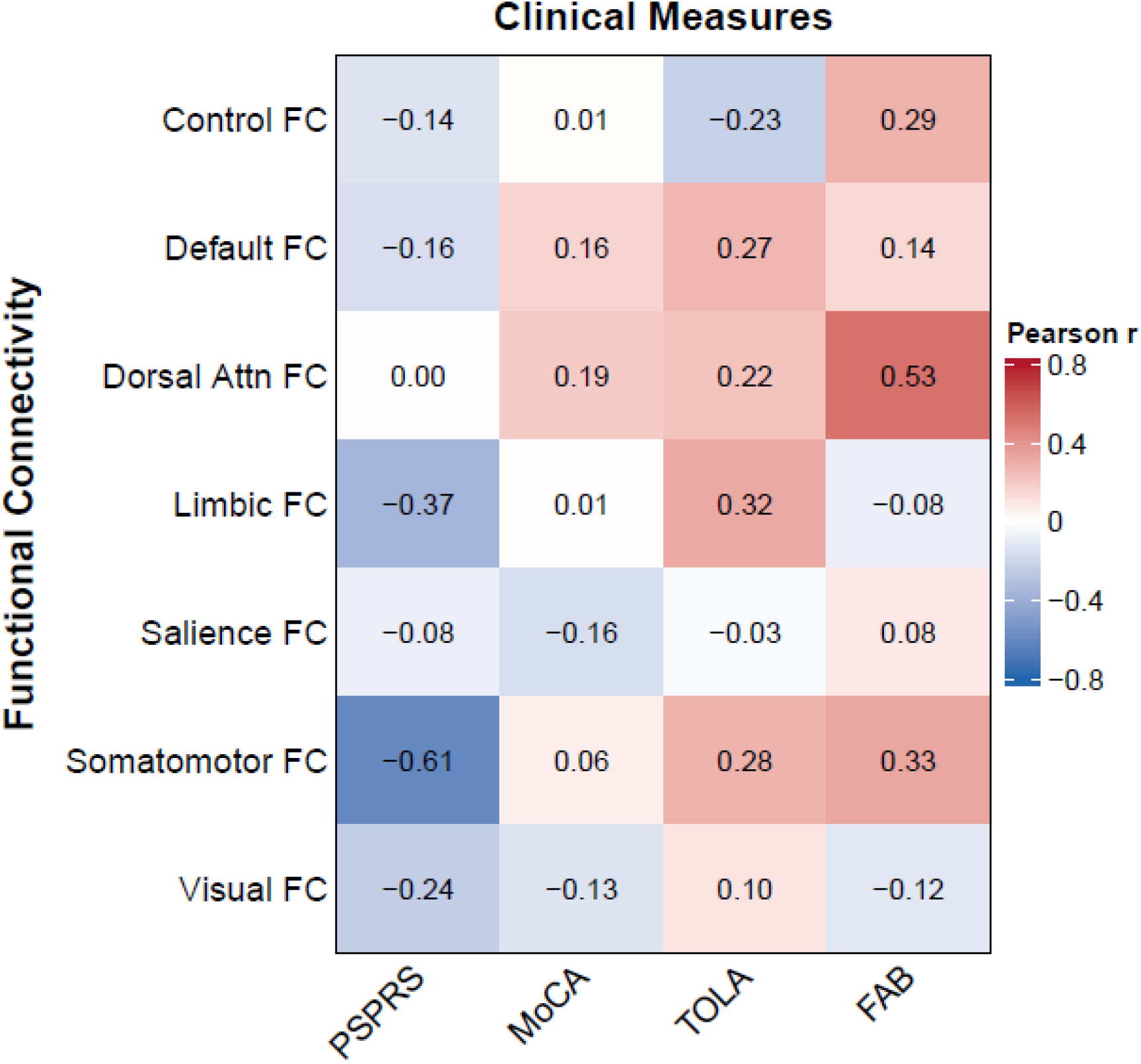
Heatmap for correlations between within-network functional connectivity and clinical measures Associations were not significant after FDR correction.

**Figure 4.**
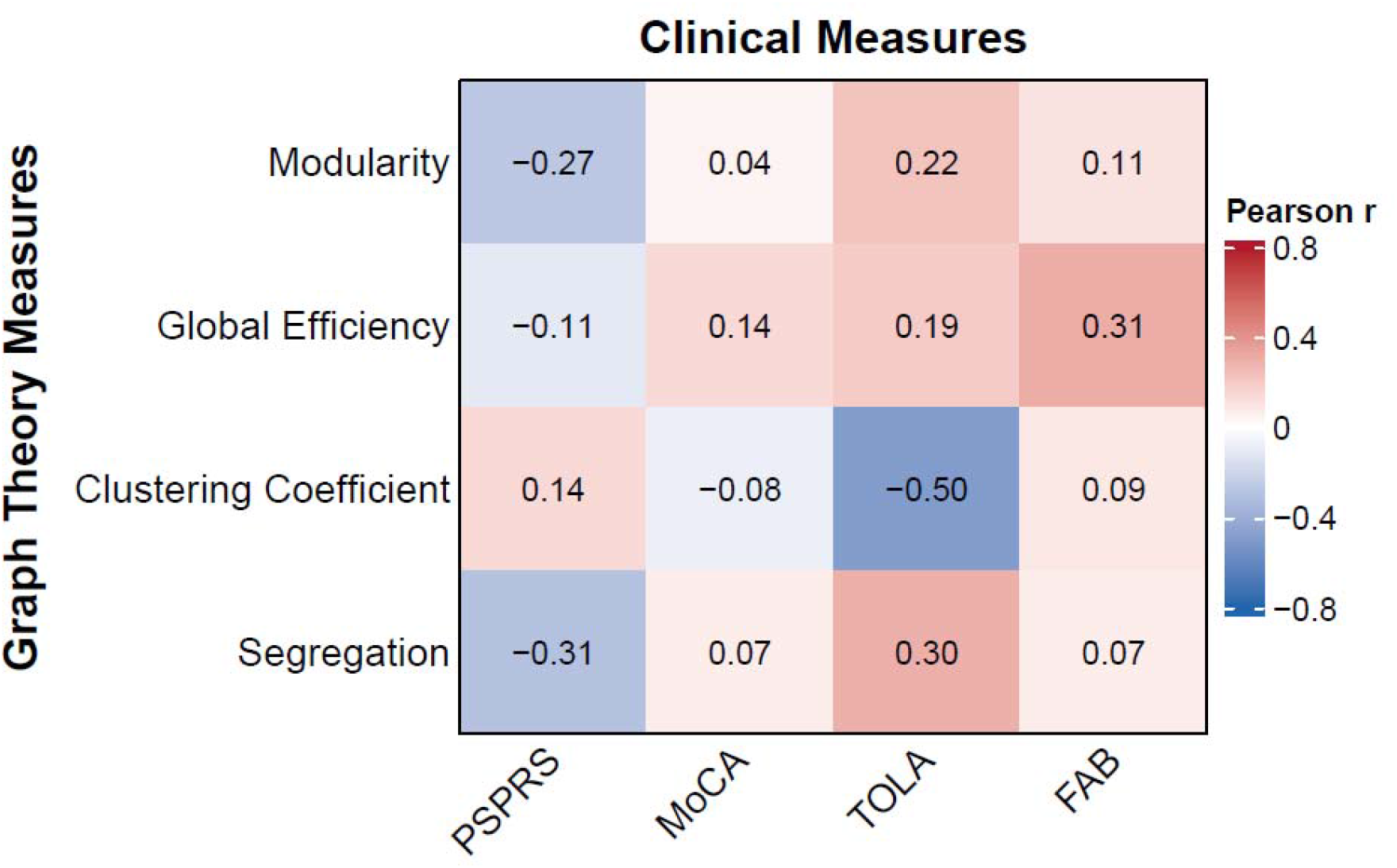
Heatmap for correlations between graph theory and clinical measures Associations were not significant after FDR correction.

**Table 2.**
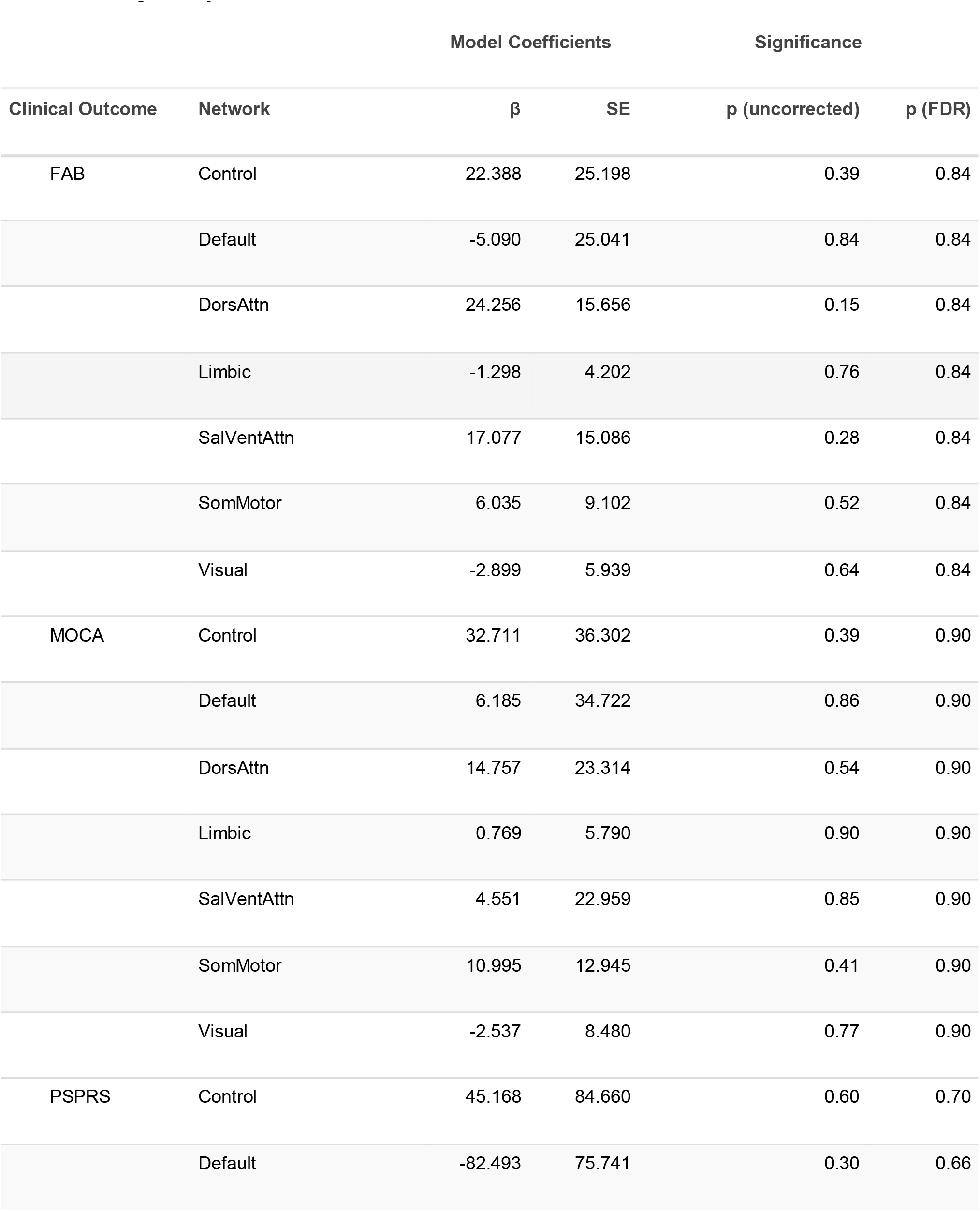
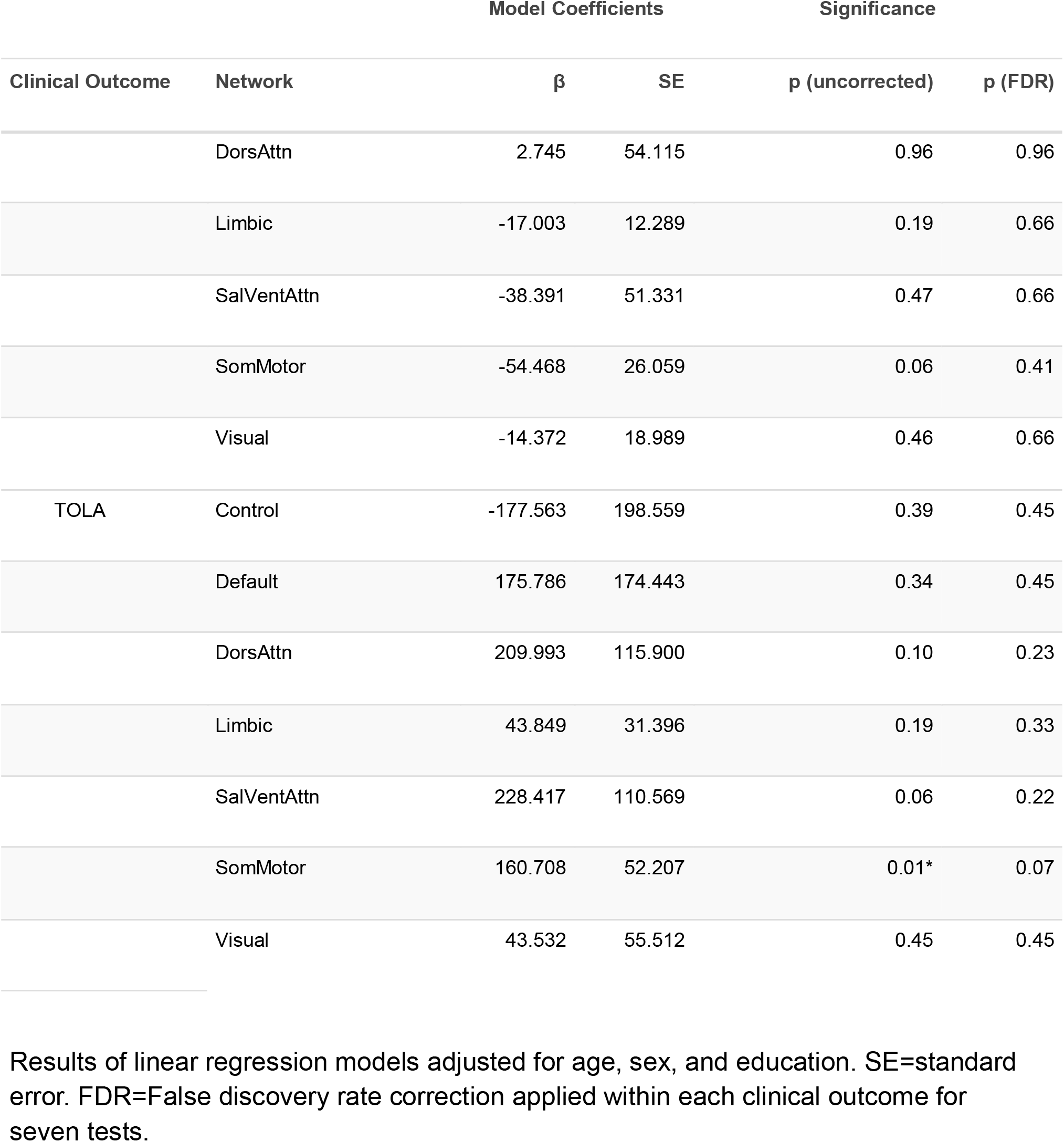
Associations between functional connectivity measures across seven networks by Yeo parcellation and clinical measures

|  |  | **Model Coefficients** |  | **Significance** |  |
| **Clinical Outcome** | **Network** | **β** | **SE** | **p (uncorrected)** | **p (FDR)** |
|---|---|---|---|---|---|
| FAB | Control | 22.388 | 25.198 | 0.39 | 0.84 |
|  | Default | -5.090 | 25.041 | 0.84 | 0.84 |
|  | DorsAttn | 24.256 | 15.656 | 0.15 | 0.84 |
|  | Limbic | -1.298 | 4.202 | 0.76 | 0.84 |
|  | SalVentAttn | 17.077 | 15.086 | 0.28 | 0.84 |
|  | SomMotor | 6.035 | 9.102 | 0.52 | 0.84 |
|  | Visual | -2.899 | 5.939 | 0.64 | 0.84 |
| MOCA | Control | 32.711 | 36.302 | 0.39 | 0.90 |
|  | Default | 6.185 | 34.722 | 0.86 | 0.90 |
|  | DorsAttn | 14.757 | 23.314 | 0.54 | 0.90 |
|  | Limbic | 0.769 | 5.790 | 0.90 | 0.90 |
|  | SalVentAttn | 4.551 | 22.959 | 0.85 | 0.90 |
|  | SomMotor | 10.995 | 12.945 | 0.41 | 0.90 |
|  | Visual | -2.537 | 8.480 | 0.77 | 0.90 |
| PSPRS | Control | 45.168 | 84.660 | 0.60 | 0.70 |
|  | Default | -82.493 | 75.741 | 0.30 | 0.66 |
|  |  | **Model Coefficients** |  | **Significance** |  |
| **Clinical Outcome** | **Network** | **β** | **SE** | **p (uncorrected)** | **p (FDR)** |
|  | DorsAttn | 2.745 | 54.115 | 0.96 | 0.96 |
|  | Limbic | -17.003 | 12.289 | 0.19 | 0.66 |
|  | SalVentAttn | -38.391 | 51.331 | 0.47 | 0.66 |
|  | SomMotor | -54.468 | 26.059 | 0.06 | 0.41 |
|  | Visual | -14.372 | 18.989 | 0.46 | 0.66 |
| TOLA | Control | -177.563 | 198.559 | 0.39 | 0.45 |
|  | Default | 175.786 | 174.443 | 0.34 | 0.45 |
|  | DorsAttn | 209.993 | 115.900 | 0.10 | 0.23 |
|  | Limbic | 43.849 | 31.396 | 0.19 | 0.33 |
|  | SalVentAttn | 228.417 | 110.569 | 0.06 | 0.22 |
|  | SomMotor | 160.708 | 52.207 | 0.01* | 0.07 |
|  | Visual | 43.532 | 55.512 | 0.45 | 0.45 |

Results of linear regression models adjusted for age, sex, and education. SE=standard error. FDR=False discovery rate correction applied within each clinical outcome for seven tests.

**Table 3.** Associations between graph theory metrics and clinical measures

|  |  | Model Coefficients |  | Significance |  |
|---|---|---|---|---|---|
| **Clinical Outcome** | **Graph Theory Metric** | **β** | **SE** | **p (uncorrected)** | **p (FDR)** |
| FAB | Cluster Coefficient | 26.170 | 36.859 | 0.49 | 0.70 |
|  | Global_Efficiency | 39.145 | 49.139 | 0.44 | 0.70 |
|  | Modularity | -10.145 | 25.726 | 0.70 | 0.70 |
| MOCA | Cluster Coefficient | 16.262 | 54.159 | 0.77 | 0.99 |
|  | Global_Efficiency | 39.215 | 72.112 | 0.60 | 0.99 |
|  | Modularity | 0.380 | 37.196 | 0.99 | 0.99 |
| PSPRS | Cluster Coefficient | 137.545 | 117.618 | 0.26 | 0.35 |
|  | Global_Efficiency | -78.719 | 165.134 | 0.64 | 0.64 |
|  | Modularity | -107.390 | 79.081 | 0.20 | 0.35 |
| TOLA | Cluster Coefficient | -503.395 | 239.699 | 0.06 | 0.15 |
|  | Global_Efficiency | 321.418 | 376.760 | 0.41 | 0.41 |
|  | Modularity | 296.366 | 173.058 | 0.11 | 0.15 |

Results of linear regression models adjusted for age, sex, and education. SE=standard error. FDR=False discovery rate correction applied within each clinical outcome for three tests.

## Graph Theory Measures

When compared to controls, PSP participants were found to have significantly lower network modularity (0.108 vs. 0.179 respectively, p<0.0001). No significant differences were found between PSP participants and controls for global efficiency and clustering coefficient (Figure 2). Graph theory measures were standardized to Z-scores to enable visual comparison across metrics with differing units and scales. No significant differences were found for any graph theory measures when comparing across PSP subtypes (Supplementary table 1). Results did not change when including total cortical volume as an additional covariate (data not shown).

Correlation analyses between clinical and graph theory measures among PSP participants showed a significant negative linear relationship between cluster coefficient and total TOLA scores (r=-0.50, p=0.018); however, this association was not significant after FDR correction (Figure 3).

## Discussion

In this study, we compared rsfMRI connectivity measures between participants with PSP and age-and sex-matched healthy controls. We also examined differences between 3 PSP clinical variants: PSP-RS, PSP-SL, and PSP-CBS. Our findings demonstrate widespread functional connectivity disruptions of resting-state functional networks in these PSP variants, with lower connectivity within most canonical large-scale networks. When examining graph theory measures, we found significantly lower network modularity among the PSP participants compared to controls, along with preserved clustering coefficients and global network efficiency. There were no significant differences in functional connectivity patterns and network organization among the three PSP subtypes. Additionally, higher functional connectivity within the somatomotor network was associated with higher TOLA scores, indicating better motor functioning. Taken together, these results suggest that network reorganization in PSP shares important commonalities across clinical variants and may reflect adaptations to ongoing neurodegeneration that prioritize preservation of global network efficiency at the expense of reduced network separation. This network de-differentiation may result from regions in less affected networks taking on the functional tasks of nodes that are in more advanced stages of network degeneration.

When comparing functional connectivity measures across canonical networks of interest, we found significant differences between PSP and controls. Specifically, these included lower connectivity in frontoparietal control, default mode, dorsal attention, somatomotor, and visual networks. The frontoparietal control network includes the dorsolateral prefrontal cortex, anterior inferior parietal lobe, and anterior cingulate cortex. Functionally it acts as a hub, sharing a large degree of connectivity to other diverse brain networks and is thought to be involved in higher cognitive functions including task adaptation, working memory, and decision-making^40^. It is extensively involved in executive function and is known to be disrupted in both neurologic and psychiatric disorders^41^. This is relevant in PSP given the high proportion (up to 74%) of patients who develop executive dysfunction (the most commonly affected cognitive domain in PSP)^42–44^. The default mode network includes the posterior cingulate cortex, medial prefrontal cortex, medial temporal lobe, and the angular gyrus. It supports social cognition, introspection, and episodic memory^45^ and has wide implications in neurodegenerative and psychiatric disorders^46–48^ This network plays a central role in internally directed cognition and is also frequently affected in neurodegenerative disorders such as AD^49^ ^50^. Reduced DMN connectivity has previously been observed in PSP and may reflect degeneration within frontal and midline cortical structures that support executive, social, and memory processes^51^. The somatomotor and dorsal attention networks are particularly relevant to the clinical features of PSP. The somatomotor network includes the primary motor cortex, primary somatosensory cortex, supplementary motor area, and extends to the superior temporal gyrus anatomically. This network is crucial in motor execution and planning, somatosensory processing, and auditory perception. The dorsal attention network has key nodes in the intraparietal sulcus and frontal eye fields and mediates voluntary, top-down deployment of visuospatial attention. PSP is characterized by postural instability, impaired voluntary eye movements, and motor dysfunction. Thus, disruption of these networks may contribute to the motor and visuospatial deficits commonly observed in PSP. Notably, our unadjusted correlational analyses suggested relationships between somatomotor connectivity and disease severity (PSPRS) and between dorsal attention connectivity and frontal executive function (FAB), though these did not survive correction for multiple comparisons, likely due to our small sample size. This suggests a need for larger studies.

Network modularity aims to quantify the connectivity within subnetworks, or modules, relative to connectivity between these subneworks^36^. Decreased modularity suggests a breakdown in the functional specialization of large-scale brain networks, or greater network de-differentiation. Our findings of lower within network functional connectivity and lower network modularity are consistent with previous studies showing that neurodegenerative diseases selectively disrupt the topological organization of brain networks. This concept of network de-differentiation is most clearly seen in the Alzheimer’s disease spectrum, where structural connectivity derangements have been observed to precede functional connectivity loss in earlier disease stages, though findings have varied^12,37^. In later stages of disease, these measures tend to universally degrade with clustering coefficient remaining relatively spared^38^. Given that most of our PSP cohort was assessed at mid-stage disease severity, our observations may point to networks that have lost intra-subnetwork connectivity but have preserved or gained inter-subnetwork connectivity with unrelated nodes, thus increasing randomness in connectivity, with a corresponding decrease in the networks’ functional specialization^39^.

To our knowledge, there have been few investigations focused on graph theory-derived functional connectivity measures in PSP. One study of similar size to ours found significantly lower connectivity between cortical and deep gray matter structures in PSP (17 PSP-RS, 4 PSP-PGF, 1 PSP-P, and 1 PSP-CBS) vs controls, with no differences in whole-brain modularity^16^. This discrepancy may in part be due to the use of different PSP variants from those in our study, different graph theory measures (path length vs. global efficiency), and different cortical parcellation techniques.

We did not find differences in network connectivity measures between PSP subtypes although we did observe a trend for lower connectivity within the ventral attention network among PSP-RS individuals compared to PSP-SL and PSP-CBS that did not survive multiple test correction. This may be explained by small and unequal numbers of PSP participants across variants, but may also reflect the generally greater attentional disruptions clinically observed in PSP-RS. The ventral attention, or salience, network mediates stimulus-driven attentional reorienting. PSP-RS is distinguished from other PSP variants by the early and prominent emergence of, postural instability, vertical gaze palsy, and frontal-executive dysfunction, the latter of which depend in part or play a role in attentional reorienting mechanisms. By contrast speech and limb praxis deficits are less dependent on attentional processes. Thus, preferential disruption of the ventral attention network in PSP-RS may reflect the disproportionate burden of attentional and oculomotor dysfunction that characterizes this subtype relative to others. Future studies with larger numbers of both PSP-RS participants and other PSP variants are needed to confirm this finding. In addition, although PSP subtypes present heterogeneously, the underlying tauopathy may affect system-wide functional connectivity to a similar extent. There are however, reported differences in connectivity between PSP variants; one study examined functional connectivity between various anatomical regions of interest and found PSP-RS had widespread network abnormalities along the dentato-thalamo-cortical pathway as well as within the frontal cortex. These abnormalities overlapped somewhat with cortical PSP (defined as either PSP-SL, PSP-CBS, or PSP-F) however with more pronounced substantia nigra involvement in PSP-RS. Subcortical PSP (either PSP-P, PSP-PGF, PSP-PI, or PSP-OM), by contrast, had abnormalities concentrated in subcortical structures as expected^14^.

This study has several notable strengths. First, we employed a comprehensive, systems-level approach using rs-fMRI to examine brain connectivity in PSP, integrating both graph theory metrics and canonical network analyses to provide complementary perspectives on network dysfunction. This dual analytic strategy allowed us to characterize both global topological properties and specific network alterations, offering a more complete picture of functional disruption in PSP than either approach alone. Additionally, we included well-characterized PSP participants with clinical diagnoses based on MDS-PSP criteria who underwent comprehensive clinical phenotyping with validated instruments (PSPRS, MoCA, FAB), enabling robust clinical-imaging correlations. By examining connectivity across seven canonical resting-state networks defined by the established Yeo atlas rather than selecting regions a priori, we avoided biasing our analysis toward specific networks and captured a comprehensive view of large-scale functional architecture.

Several limitations should be considered when interpreting our findings. First, our study lacked autopsy confirmation of PSP diagnosis, relying instead on clinical diagnostic criteria. While all participants met MDS criteria for probable or possible PSP, pathological heterogeneity cannot be excluded, and some participants may have alternative or mixed underlying pathologies. Second, our sample size was relatively small (n=23), which may have limited our power to detect differences between PSP clinical variants and identify more subtle network alterations. The uneven distribution across subtypes (RS=11, SL=7, CBS=5) and lack of inclusion of other PSP subtypes further constrained subtype-specific analyses. Third, we focused on rs-fMRI connectivity measures and did not incorporate additional biomarkers such as CSF tau species, neurofilament light chain, or tau PET imaging, which could provide complementary information about disease pathophysiology and staging.

## Conclusion

Our study demonstrates that PSP is associated with widespread disruption of intrinsic functional brain networks, characterized by preserved global network efficiency and significantly reduced modularity and segregation. This may reflect network dedifferentiation as an adaptive mechanism in the face of continued loss of synaptic connections. In addition, we found decreased connectivity across multiple canonical resting-state networks including frontoparietal control, default, dorsal attention, somatomotor, and visual networks. Importantly, these network-level alterations appear broadly similar across three PSP clinical variants investigated in this study. Our findings support the concept that PSP represents a distributed network disorder with unifying pathophysiological mechanisms across heterogeneous clinical presentations. If validated in larger cohorts, these findings would support rs-fMRI metrics as potential biomarkers for PSP that capture disease-related network dysfunction independently of clinical phenotype. Future directions should include longitudinal studies to characterize the temporal evolution of network disruption in relation to clinical progression.

## Data Availability

All data produced in the present study are available upon reasonable request to the authors

## Acknowledgments

We thank our PSP and Biocard study participants for providing the clinical and imaging data for this study. This work was supported by NIH/NIA grant AG059891 (Pantelyat).

---

**Supplementary Table 1.** Comparisons of within-network functional connectivity and graph theory measures across PSP variants

|  | PSP-RS | PSP-SL | PSP-CBS | P (uncorrected) | p (FDR) |
|---|---|---|---|---|---|
| Control | 0.070 | 0.09 | 0.09 | 0.25 | 0.86 |
| Default | 0.12 | 0.12 | 0.17 | 0.99 | 0.99 |
| DorsAttn | 0.101 | 0.126 | 0.138 | 0.61 | 0.86 |
| Limbic | 0.362 | 0.256 | 0.267 | 0.56 | 0.87 |
| SalVentAttn | 0.114 | 0.132 | 0.204 | **0.03** | 0.18 |
| SomMotor | 0.221 | 0.230 | 0.225 | 0.99 | 0.99 |
| Visual | 0.441 | 0.403 | 0.329 | 0.45 | 0.86 |
| Modularity | 0.110 | 0.111 | 0.111 | 0.94 | 0.94 |
| Cluster Coefficient | 0.090 | 0.103 | 0.102 | 0.36 | 0.55 |
| Global Efficiency | 0.752 | 0.743 | 0.760 | 0.25 | 0.55 |

## Notes

### Competing Interest Statement

The authors have declared no competing interest.

### Author Declarations

IRB of Johns Hopkins Medicine gave ethical approval for this work.

